# Case Series of Thrombosis with Thrombocytopenia Syndrome following COVID-19 vaccination—United States, December 2020–August 2021

**DOI:** 10.1101/2021.11.10.21266063

**Authors:** Isaac See, Allison Lale, Paige Marquez, Michael B. Streiff, Allison P. Wheeler, Naomi K. Tepper, Emily Jane Woo, Karen R. Broder, Kathryn M. Edwards, Ruth Gallego, Andrew I. Geller, Kelly A. Jackson, Shashi Sharma, Kawsar R. Talaat, Emmanuel B. Walter, Imo J. Akpan, Thomas L. Ortel, Shannon C. Walker, Jennifer C. Yui, Tom T. Shimabukuro, Adamma Mba-Jonas, John R. Su, David K. Shay

## Abstract

**Background:** Thrombosis with thrombocytopenia syndrome (TTS) is a potentially life-threatening condition associated with adenoviral-vectored COVID-19 vaccination. TTS presents similarly to autoimmune heparin-induced thrombocytopenia. Twelve cases of cerebral venous sinus thrombosis following Janssen/Johnson & Johnson (Ad26.COV2.S) COVID-19 vaccination have been described.

**Objective:** Describe surveillance data and reporting rates of TTS cases following COVID-19 vaccination.

**Design:** Case series.

**Setting:** United States

**Patients:** Case-patients reported to the Vaccine Adverse Event Reporting System (VAERS) receiving COVID-19 vaccine from December 14, 2020 through August 31, 2021, with thrombocytopenia and thrombosis (excluding isolated ischemic stroke or myocardial infarction). If thrombosis was only in an extremity vein or pulmonary embolism, a positive enzyme-linked immunosorbent assay for anti-platelet factor 4 antibody was required.

**Measurements:** Reporting rates (cases/million vaccine doses) and descriptive epidemiology.

**Results:** 52 TTS cases were confirmed following Ad26.COV2.S (n=50) or mRNA-based COVID-19 (n=2) vaccination. TTS reporting rates were 3.55 per million (Ad26.COV2.S) and 0.0057 per million (mRNA-based COVID-19 vaccines). Median age of patients with TTS following Ad26.COV2.S vaccination was 43.5 years (range: 18–70); 70% were female. Both TTS cases following mRNA-based COVID-19 vaccination occurred in males aged >50 years. All cases following Ad26.COV2.S vaccination involved hospitalization including 32 (64%) with intensive care unit admission. Outcomes of hospitalizations following Ad26.COV2.S vaccination included death (12%), discharge to post-acute care (16%), and discharge home (72%).

**Limitations:** Under-reporting and incomplete case follow-up.

**Conclusion:** TTS is a rare but serious adverse event associated with Ad26.COV2.S vaccination. The different demographic characteristics of the two cases reported after mRNA-based COVID-19 vaccines and the much lower reporting rate suggest that these cases represent a background rate.

**Funding Source:** CDC

## Introduction

Thrombosis with thrombocytopenia syndrome (TTS), also referred to as vaccine-induced thrombotic thrombocytopenia (VITT), is a rare condition recently described among persons who received the adenoviral-vectored COVID-19 vaccines made by Janssen/Johnson & Johnson (Ad26.COV2.S) and Oxford/AstraZeneca (ChAdOx1 nCoV-19) [1-5]. Initial recognition of the condition precipitated a pause in the administration of Ad26.COV2.S in the United States and modifications of ChAdOx1 nCoV-19 vaccine policy in other countries [6-7]. Based on the distinctive clinical and laboratory features of the syndrome, epidemiologic clustering of timing after receipt of adenoviral-vectored COVID-19 vaccines, and plausible pathogenic mechanisms, causal associations between receipt of adenoviral vectored vaccines and TTS are likely [8].

Reported rates of TTS after ChAdOx1 nCoV-19, which is not approved or authorized for use in the United States, range from 13–39 cases per million vaccine doses administered, depending on the publication [2,9-11]. The first reports of TTS following ChAdOx1 nCoV-19 described a high case-fatality rate and features shared with autoimmune or spontaneous heparin-induced thrombocytopenia (HIT) [1-3], including thrombocytopenia, thrombosis, and the presence of antibodies to platelet factor 4 (PF4) [1-3,12] in the absence of prior exposure to heparin. These reports described positive results for antibody to PF4 measured by enzyme-linked immunosorbent assay (ELISA) and in many cases, positive results for various functional HIT platelet assays, which evaluate the ability of anti-PF4 antibodies to activate platelets and cause thrombosis [1-3,13]. Preliminary hypotheses on the pathogenesis of TTS generally posit mechanisms related to the adenovirus vector platform, including triggering of HIT-like antibodies from adenoviral DNA, from cell-line-derived factors, or from splice variants of the SARS-CoV-2 spike protein caused by the adenoviral vector platform [1,14-15].

TTS after ChAdOx1 nCoV-19 vaccine receipt has been described in detail [16]. In the United States, the first 12 cases of TTS following Ad26.COV2.S involving cerebral venous sinus thrombosis were described in a preliminary report [5]; and additional case reports have been published including one case of TTS following mRNA COVID-19 vaccination [17].

This report is the first comprehensive review of TTS following COVID-19 vaccination in the United States. We describe epidemiology and clinical characteristics of the case reports to inform clinicians and public health officials.

## Methods

### Data Sources

The Vaccine Adverse Events Reporting System (VAERS) is a passive surveillance (spontaneous reporting) system for adverse events following immunization jointly administered by the U.S. Centers for Disease Control and Prevention (CDC) and the Food and Drug Administration (FDA) [18]. VAERS accepts all reports of possible adverse events following vaccination from patients, clinicians, vaccine manufacturers, and the public, regardless of clinical severity of the event or its causal association with vaccination. The VAERS database was searched for potential reports of TTS received during December 14, 2020 through August 31, 2021, in the following manner: 1) on April 9, 2021, a retrospective search was conducted for reports received during December 14, 2020 through April 8, 2021; 2) prospective searches were conducted beginning on April 9, 2021. All data for this report were retrieved by August 31, 2021. Reports were screened for Medical Dictionary for Regulatory Activities (MedDRA) standardized codes (Preferred Terms [PTs]) [19] assigned to reports by professional medical coders, as well as language in the symptom or laboratory text fields of the report, that described symptoms, laboratory values, and medical conditions that could indicate TTS (see Supplemental Methods).

Vaccine administration data were reported by public health jurisdictions to CDC (Supplemental Methods) [20].

### Case definitions

We used the following case definitions for TTS. A Tier 1 case had thrombosis in an unusual location for a thrombus (i.e., cerebral vein, visceral artery or vein, extremity artery, central artery or vein) and new-onset thrombocytopenia (i.e., platelet count <150,000/µL) occurring any time after receipt of a COVID-19 vaccine. Tier 2 cases had new-onset thrombocytopenia, thrombosis in an extremity vein or pulmonary artery in the absence of thrombosis at a Tier 1 location, and a positive anti-PF4 antibody ELISA test or functional HIT platelet test occurring any time after receipt of COVID-19 vaccine. Given the very unusual clinical presentation of the Tier 1 cases, additional laboratory abnormalities were not required, whereas the inclusion of positive HIT testing results for cases with only common thromboses (Tier 2) was intended to limit the number of potential false positive cases.

Confirmation of case status required review of patient data from medical records and in some cases also discussion with providers. All cases were also reviewed with clinicians from CDC’s Clinical Immunization Safety Assessment (CISA) Project (Supplemental Methods) [21].

### Data analysis

TTS reporting rates were calculated per million doses of Ad26.COV2.S vaccine administered and stratified by sex and age group. Overall reporting rates were calculated per million mRNA COVID-19 vaccine (i.e., Moderna and Pfizer-BioNTech) doses administered. Booster dose administration was excluded from reporting rate calculations.

Data were collected and managed using Research Electronic Data Capture (REDCap) electronic data capture tools hosted at CDC. REDCap is a secure, web-based software platform designed to support data entry and management [22,23]. Descriptive analysis of demographic and clinical data associated with TTS cases was conducted. Demographics, risk factors for thrombosis, platelet nadir, and timing of clinical presentation were compared between Tier 1 and Tier 2 cases using chi-square or Fisher’s exact test, as appropriate, for categorical variables and using Wilcoxon rank-sum test for continuous variables. To permit comparisons with other reports concerning TTS, supplemental results describe whether cases met definite or probable VITT criteria established by the United Kingdom (UK) Expert Hematology Panel [17]. Venous thromboembolism risk factors were assessed according to published literature [24-25]. Data analyses were conducted with SAS 9.4 (SAS Institute, Cary, NC).

### Human subjects

This activity was reviewed by the CDC Human Subjects Office, determined to be a non-research public health surveillance project, and was conducted consistent with applicable federal law and policy (see e.g., 45 C.F.R. part 46.102(I)(2), 21 C.F.R. part 56; 42 U.S.C. §241(d); 5 U.S.C. §552a; 44 U.S.C. §3501 et seq).

## Results

### TTS reporting rates

Of 579,301 total reports to VAERS after COVID-19 vaccination during the analytic period, 1,122 were identified as potential cases based on initial screening of VAERS reports. Of these, 51 are pending adjudication, 6 had insufficient information was contained in the report to determine (e.g., the report included information about thrombosis only and no provider listed to obtain further information) and 953 did not meet case definition criteria. Of the 112 remaining reports, confirmed to have thrombosis and thrombocytopenia after COVID-19 (i.e., potentially meeting criteria for the TTS case definition), 66 reports were excluded because they were duplicate reports (n=30); active COVID-19 infection was documented at the time of TTS diagnosis (n=5); thrombocytopenia was not new-onset (n=2); TTS symptoms preceded COVID-19 vaccination (n=3); thrombocytopenia was most likely artifactual (n=1); or thrombosis was more likely due to another cause (n=19). After these exclusions, 52 reports were determined to meet the CDC case definition for TTS: 50 cases after Ad26.COV2.S and 2 after mRNA COVID-19 vaccination (Figure 1).

**Figure 1:**
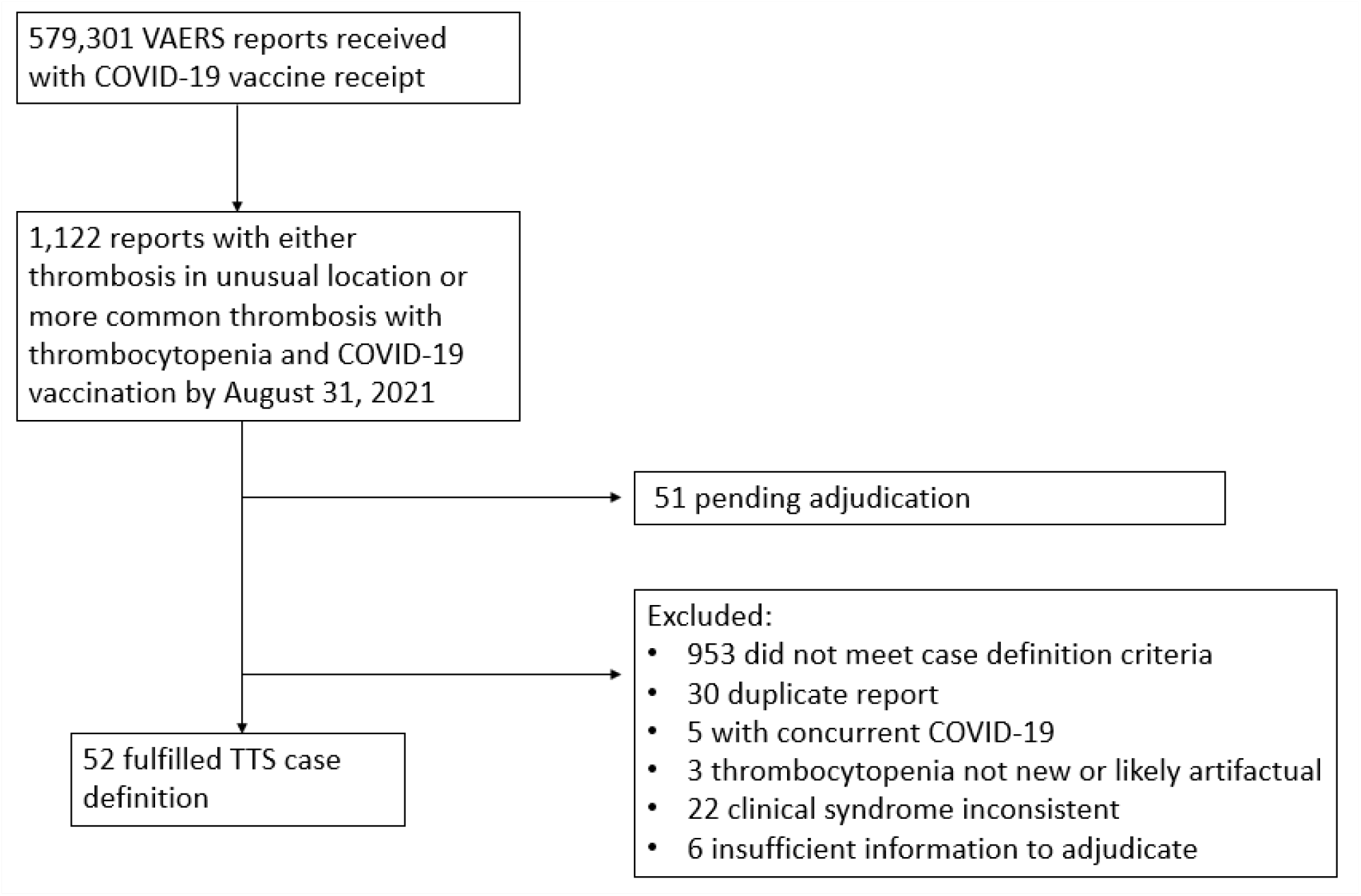
Reports to the Vaccine Adverse Event Reporting System (VAERS) confirmed to be thrombosis with thrombocytopenia syndrome (TTS), United States, December 2020–August 2021

Given administration of 14.1 million doses of Ad26.COV2.S and 351 million doses of mRNA COVID-19 vaccines, the overall TTS reporting rate (per million vaccine doses administered) was 3.55 after Ad26.COV2.S and 0.0057 after mRNA COVID-19 vaccination. Reporting rates were highest following receipt of Ad26.COV2.S vaccine among females aged 30-39 years (10.6 per million doses) and 40-49 years (8.12 per million doses) (Table 1).

**Table 1:** Reporting rates for thrombosis with thrombocytopenia syndrome (TTS) following receipt of Ad26.COV2.S (Janssen/Johnson & Johnson COVID-19) and mRNA COVID-19 vaccines (Pfizer-BioNTech BNR162b2 or Moderna mRNA-1263)—United States, December 2020–August 2021

| **Reporting rate of TTS after Ad26.COV2.S vaccination (cases per million doses administered)** |  |  |
| --- | --- | --- |
| **Overall** | 3.55 |  |
| **Age group (years)** | **Female** | **Male** |
| 0-17* | 0 | 0 |
| 18-29 | 4.59 | 1.92 |
| 30-39 | 10.6 | 1.39 |
| 40-49 | 8.12 | 4.31 |
| 50-64 | 3.99 | 1.71 |
| >65 | 1.82 | 0 |
| **Reporting rate of TTS after mRNA COVID-19 vaccines (cases per million doses administered)** |  |  |
| **Overall** | 0.0057 |  |

\* Of the 3 vaccines available during the analytic period, only the Pfizer-BioNTech COVID-19 Vaccine was authorized for use in persons aged <18 years

### Demographics, location of thrombosis, treatment, and outcomes of TTS cases following Ad26.COV2.S vaccination

The 50 TTS cases reported following Ad26.COV2.S vaccination occurred in patients with median age of 43.5 years (range: 18-70; interquartile range 36-52); 35 (70%) were female and 37 (74%) were white (Table 2). Forty-two cases were Tier 1 (84%) and 8 (16%) were Tier 2. Either a venous thrombosis or thromboembolism was found in 49/50 cases (98%). The most common anatomic region for thrombosis was cerebral venous sinuses (n=27; 54%). Eight TTS cases (16%) had arterial thrombosis (including 7 also with a venous thrombosis or thromboembolism). Thrombosis was present in more than one anatomic region for 36 cases (52%). A recognized risk factor for venous thromboembolism [24-25] was present in 33 (66%) of cases, most commonly obesity (n=23; 46%). No TTS cases occurred in females who were pregnant or within 12 weeks postpartum, or in patients with known thrombophilia. Seven cases (14%) occurred in patients with prior SARS-CoV-2 infection; demographics, presence of risk factors for venous thrombosis, and timing of illness did not differ significantly between cases with cerebral venous sinus thrombosis and other cases (Supplemental Table 1).

**Table 2:**
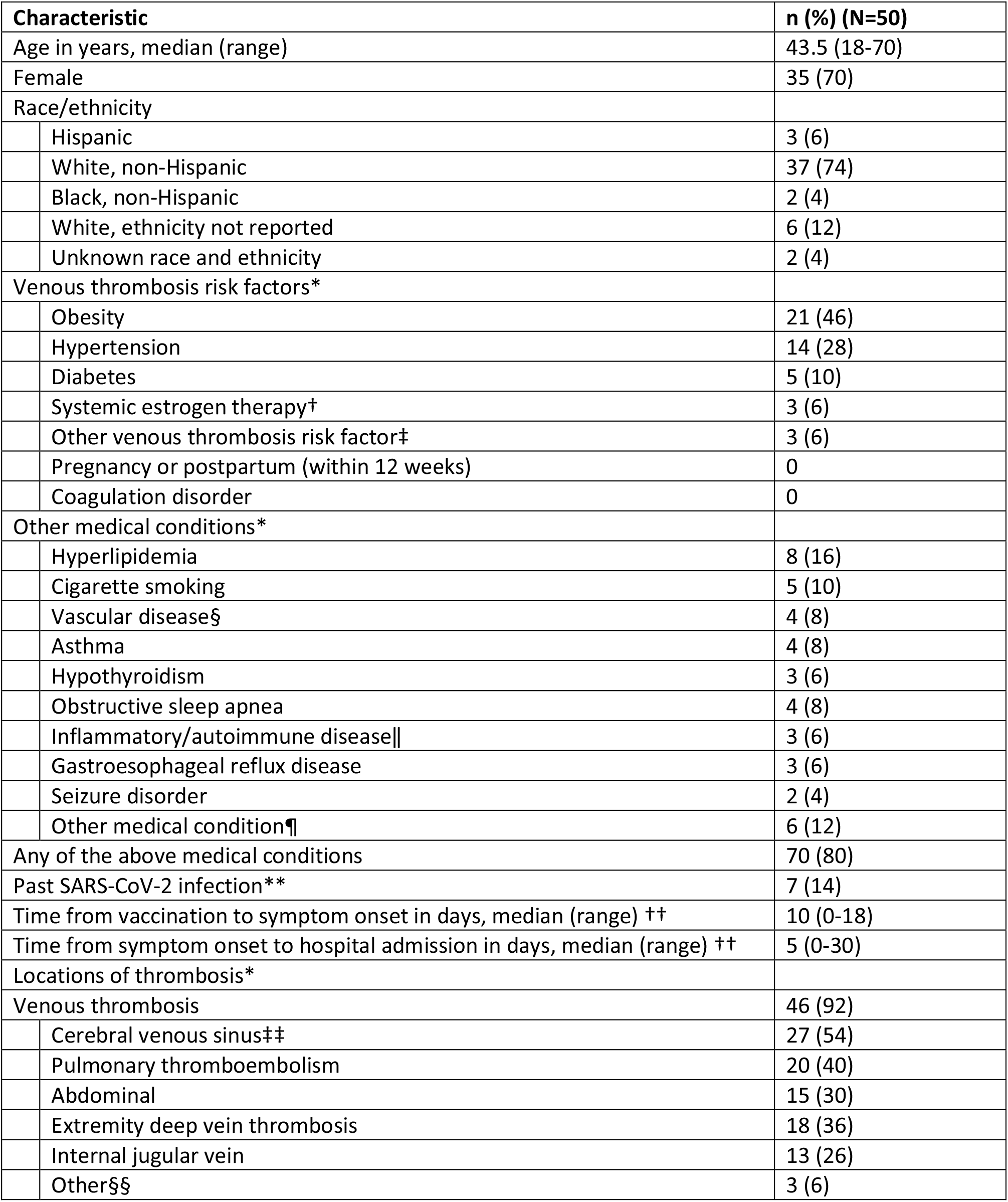
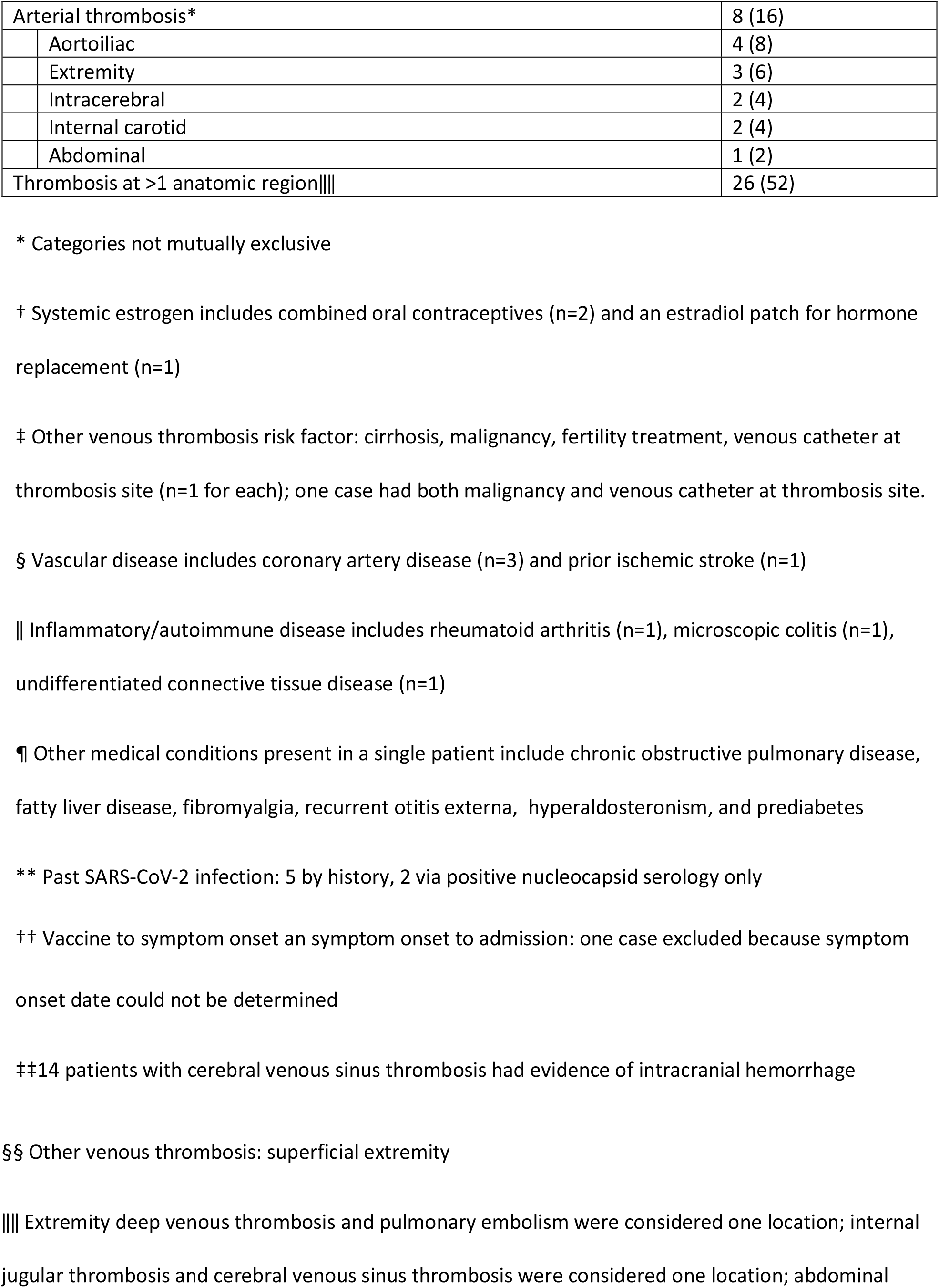
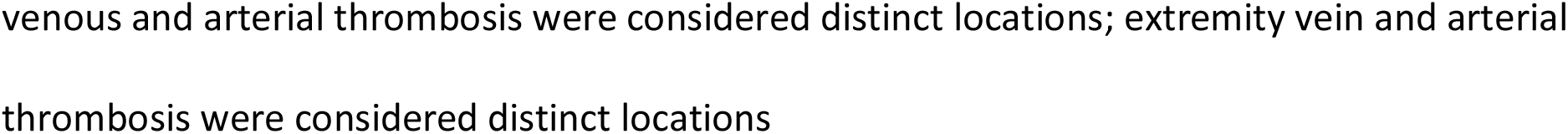
Characteristics of persons with thrombosis with thrombocytopenia syndrome reported following receipt of Ad26.COV2.S (Janssen/Johnson & Johnson COVID-19 Vaccine)—United States, December 2020–August 2021

Heparin was administered to manage thrombosis in 11/13 cases (85%) presenting before the pause in use of Ad26.COV2.S and 10/37 (27%) of the cases presenting after. Other treatments used to manage TTS were non-heparin anticoagulants (n=46; 92%), intravenous immunoglobulin (n=33; 66%), corticosteroids (n=21; 42%), platelet transfusion (n=18; 36%), and plasmapheresis (n=3; 6%). A surgical or radiologic procedure was performed in 20 cases (40%) to manage thrombosis or suspected complication, of which the most common was thrombectomy or catheter-directed thrombolysis (n=15; 30%).

All patients were hospitalized; 32 (64%) were admitted to an intensive care unit (ICU). Outcomes of the initial hospitalization were discharge home (n=36; 72%), discharge to post-acute care facility (n=8; 16%), and death (n=6; 12%). For patients discharged from the hospital alive, median hospital length of stay was 6 days (range: 1-132 days) (Supplemental Figure 1).

All sixr deaths occurred in non-Hispanic white patients with cerebral venous sinus thrombosis; five had superior sagittal sinus thrombosis. Four decedents were female; ages ranged from 29-52 years (median: 41 years). Four of the decedents had underlying medical conditions (n=4 with a risk factor for venous thromboembolism [obesity]; n=1 with asthma; n=1 with hyperlipidemia; n=1 with seizure disorder). None had documentation of prior SARS-CoV-2 infection. None received heparin. All had cerebral hemorrhage and evidence of mass effect on initial neuroimaging and died within 2 days of presentation. Median platelet count nadir among decedents was 15,500/µL (range: 9,000-44,000/µL). Anti-PF4 antibody ELISA testing was performed for four decedents; all tests were positive with optical density values >2.0.

### Laboratory findings and clinical presentation of TTS cases following Ad26.COV2.S vaccination

Symptoms began a median of 10 days after vaccination (range: 0-18 days) (Table 2; Supplemental Table 2). The most common initial symptom reported among cases was headache (n=35; 70%); headache was an initial symptom for 24/27 (89%) of cases with cerebral venous sinus thrombosis and for 11/23 (48%) of cases without cerebral venous sinus thrombosis (Supplemental table 2). Nineteen (38%) patients had at least one prior healthcare encounter for evaluation of symptoms with discharge home prior to a hospital admission and TTS diagnosis; of these, 9 were documented to have thrombocytopenia during the initial encounter (i.e., before hospital admission). In 7 cases, radiologic evaluation on initial presentation was negative for thrombosis, including 5 (of whom one later died) with initial radiologic examination negative at a site where thrombosis was later diagnosed (e.g., negative initial computed tomography venogram of the head in a patient later found to have cerebral venous sinus thrombosis on imaging).

Thrombocytopenia was present during TTS hospitalization with the exception of one case of pulmonary embolism in which thrombocytopenia was found only at the first emergency department visit following Ad26.COV2.S vaccination. The platelet count had returned to normal by the time the patient was hospitalized 6 days later with pulmonary embolism and a positive anti-PF4 antibody ELISA test.

The median platelet nadir was 32,500/µL (range: 5,000-127,000) (Table 4; Supplemental Figure 3). Platelet nadirs differed by clinical characteristics and course of disease. All patients with hospital length of stay >20 days (n=6) had platelet count nadirs <25,000/µL (Supplemental figure 4). All patients who were admitted to the hospital >12 days after symptom onset (n=6) had platelet nadirs >75,000/µL (Supplemental figure 5). Median platelet nadir for cases admitted to the ICU was 27,500/µL (range: 5,000-127,000/µL) vs 44,000/µL for cases not admitted to the ICU (range: 8,000-112,000/µL). For 39 patients discharged from the hospital alive with discharge platelet counts available, median platelet count at discharge was 167,000/µL (range: 91,000-469,000/µL) (Table 4). Of these patients, 16 (41%) had platelet count <150,000/µL at discharge.

**Table 3:**
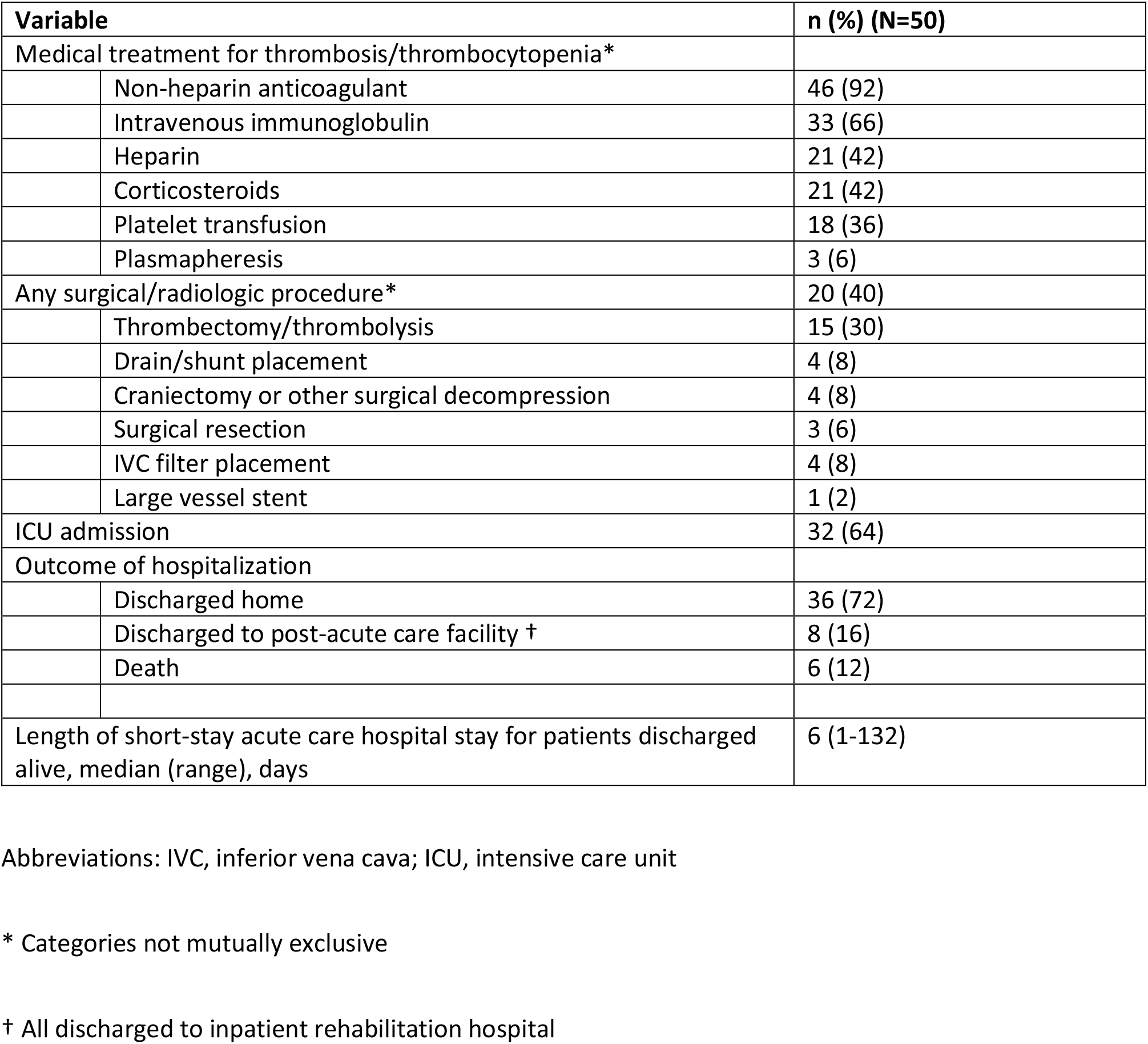
In-hospital treatment and outcomes following hospitalization for thrombosis with thrombocytopenia syndrome cases following receipt of Ad26.COV2.S (Janssen/Johnson & Johnson COVID-19 Vaccine)—United States, December 2020–August 2021

| Variable |  | n (%) (N=50) |
| --- | --- | --- |
| Medical treatment for thrombosis/thrombocytopenia* |  |  |
|  | Non-heparin anticoagulant | 46 (92) |
|  | Intravenous immunoglobulin | 33 (66) |
|  | Heparin | 21 (42) |
|  | Corticosteroids | 21 (42) |
|  | Platelet transfusion | 18 (36) |
|  | Plasmapheresis | 3 (6) |
| Any surgical/radiologic procedure* |  | 20 (40) |
|  | Thrombectomy/thrombolysis | 15 (30) |
|  | Drain/shunt placement | 4 (8) |
|  | Craniectomy or other surgical decompression | 4 (8) |
|  | Surgical resection | 3 (6) |
|  | IVC filter placement | 4 (8) |
|  | Large vessel stent | 1 (2) |
| ICU admission |  | 32 (64) |
| Outcome of hospitalization |  |  |
|  | Discharged home | 36 (72) |
|  | Discharged to post-acute care facility † | 8 (16) |
|  | Death | 6 (12) |
| Length of short-stay acute care hospital stay for patients discharged alive, median (range), days |  | 6 (1-132) |

Abbreviations: IVC, inferior vena cava; ICU, intensive care unit
\* Categories not mutually exclusive
† All discharged to inpatient rehabilitation hospital

**Table 4:**
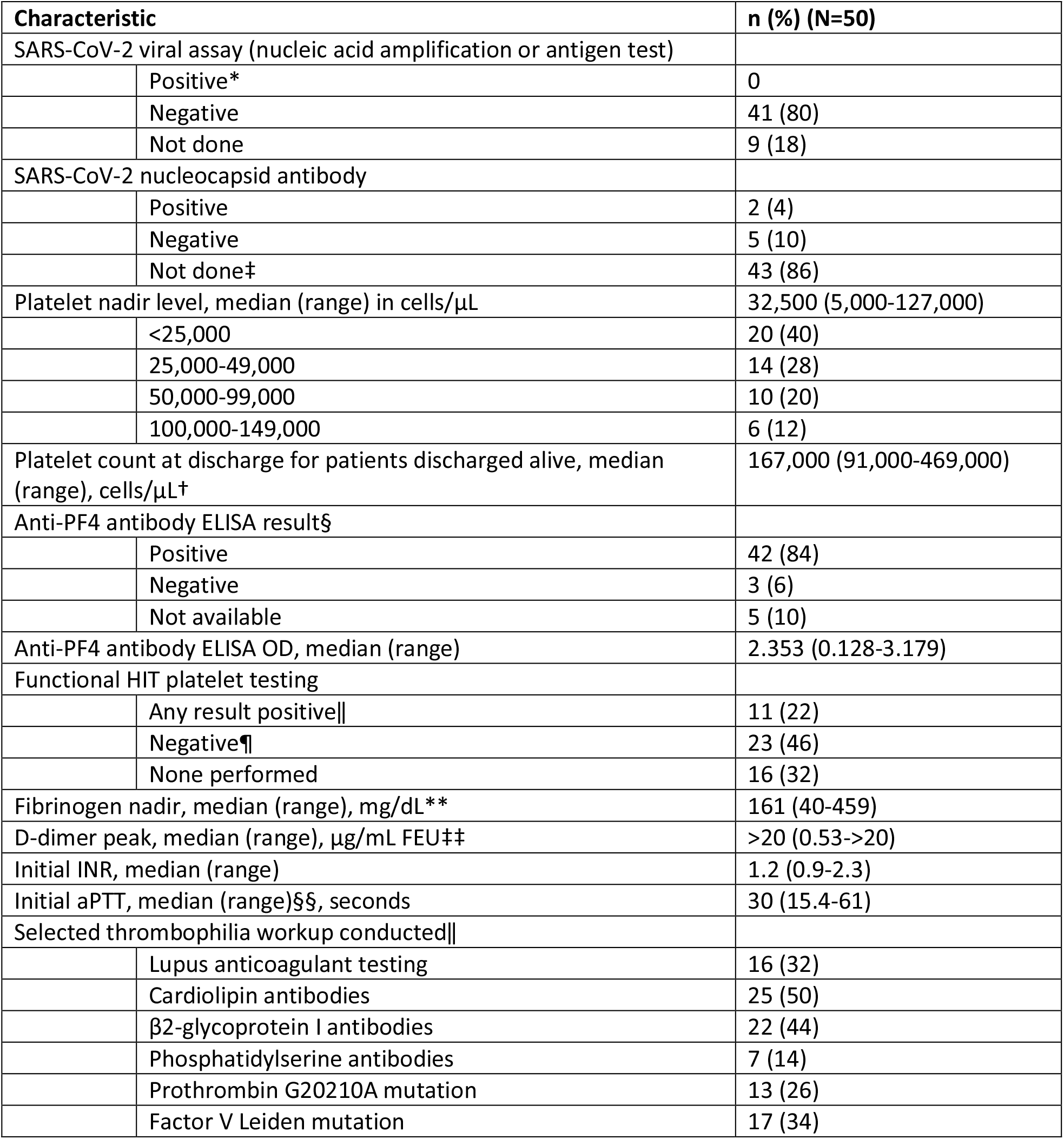
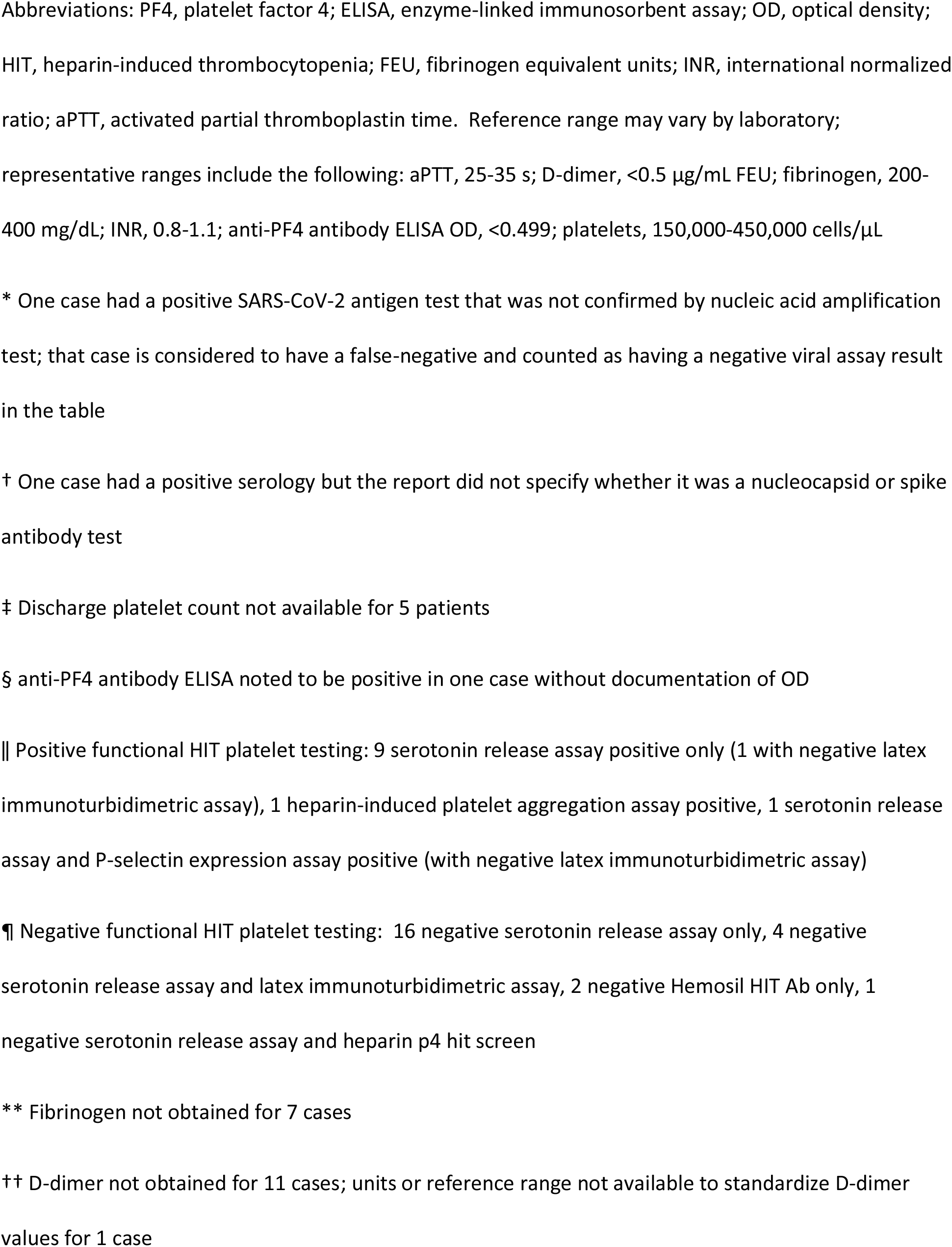
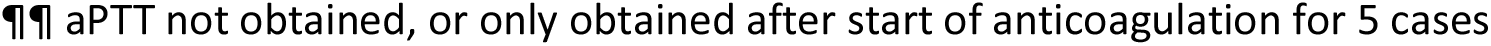
Laboratory results obtained during initial hospitalizations among persons with thrombosis with thrombocytopenia syndrome following receipt of Ad26.COV2.S (Janssen/Johnson & Johnson COVID-19 Vaccine)—United States, December 2020–August 2021

Anti-PF4 antibody ELISA results were positive in 42 (84%), negative in 3 (6%), and not performed in 5 (10%) cases (Table 4). Functional HIT platelet testing results were available for 34 and positive in 11. Of the three TTS cases following Ad26.COV2.S vaccination with negative anti-PF4 antibody ELISA results, two occurred in male patients; two were Hispanic or non-white race. All had underlying medical conditions, including hyperlipidemia (n=2) and prior vascular disease (n=2; history of myocardial infarction or ischemic stroke). All had platelet nadir >100,000/µL. Two had cerebral venous sinus thrombosis and one had abdominal vein thrombosis.

A thrombophilia evaluation (including testing for one or more of the following: lupus anticoagulant; cardiolipin, β_2_-glycoprotein I, phosphatidylserine antibodies; prothrombin G20210A mutation; Factor V Leiden mutation) was reported for 30 cases (60%); results for these 30 did not suggest an alternative cause for thrombosis (Table 4).

Supplemental Results describe evaluation of U.S. TTS cases following Ad26.COV2.S vaccination using the U.K. Expert Hematology Panel VITT case definition.

### Description of TTS cases following mRNA COVID-19 vaccination

Two TTS cases were identified following mRNA COVID-19 vaccination, both after dose 2 of the Moderna COVID-19 Vaccine. Both occurred in males aged over 50 years who were either non-white or Hispanic and had cardiovascular risk factors (hyperlipidemia and hypertension in both; diabetes in one). Both had cerebral venous sinus thrombosis, and one also had deep vein thrombosis and pulmonary emboli. Symptom onset occurred 2 and 11 days after vaccination. Platelet nadirs were <25,000/µL for one case and between 50,000-100,000/µL for the other. Anti-PF4 antibody ELISA results were positive with optical density values >2.0. One patient received intensive care and died during hospitalization; the other was discharged home.

## Discussion

This case series provides further evidence that TTS is a rare but serious condition causally associated with receipt of the Ad26.COV2.S vaccine. TTS is characterized predominantly by cerebral venous sinus thrombosis or venous thromboembolism with thrombocytopenia. Continued vigilance for TTS in patients with a history of Ad26.COV2.S vaccination, thrombocytopenia, and compatible symptoms is needed to ensure proper evaluation (e.g., anti-PF4 antibody ELISA testing) and treatment (e.g., use of non-heparin anticoagulation) of these patients [26]. Additionally, this case series provides information to contextualize two reported TTS cases following mRNA COVID-19 vaccination. Both cases were men with cardiovascular risk factors. The different demographic characteristics for the two cases and the much lower reporting rate for TTS following mRNA-based COVID-19 vaccines suggest that these 2 cases represent background cases or a different pathogenesis than the cases associated with AD26.COV2.S vaccine.

This case series highlights important clinical characteristics of TTS following Ad26.COV2.S vaccination. For example, although arterial thrombosis does occur and there are case reports of TTS involving only arterial thrombosis [27-28], TTS following Ad26.COV2.S vaccination is largely a syndrome of venous thrombosis or thromboembolism. The frequency of ICU admission, need for procedures to address thrombosis or complications, long length of stay, and fatalities confirm that TTS occurring after Ad26.COV2.S vaccination is a serious condition. The case-fatality presented in this U.S. series (12% for cases following Ad26.COV2.S vaccination) is lower than the 22% case-fatality for TTS following ChAdOx1 nCoV-19 vaccination described from the UK [16]. We found higher rates of TTS among females after Ad26.COV2.S, whereas no male/female predominance was seen in a large UK series of TTS following ChAdOx1 nCoV-19 vaccination [16]. Consistent with UK data is our finding among TTS cases following Ad26.COV2.S vaccination, the platelet count nadir may be a marker of severity or how quickly symptoms progress.

The diagnosis of TTS can be challenging. A negative initial radiologic examination in our cases did not preclude later development of thrombosis in the same location. Repeated imaging may be needed for patients who present more than once with persistent or progressive signs and symptoms of TTS. Headache was the most common initial symptom not only for patients with cerebral venous sinus thrombosis but also extracranial thrombosis, with onset usually beginning several days after vaccination. In patients with prolonged or persistent headache and thrombocytopenia following Ad26.COV2.S vaccination, search for extracranial sites of thrombosis should be considered. Some investigators recommend treatment for TTS in patients with severe headache, thrombocytopenia, elevated D-dimers, and positive anti-PF4 antibody ELISA tests after ChAdOx1 nCoV-19 vaccination, even in the absence of documented thrombosis [29].

The TTS reporting rate from U.S. VAERS data following Ad26.COV2.S vaccination (3.55 cases/million doses) is lower than rates (13-39 cases/million doses) reported for TTS following ChAdOx1 nCoV-19 vaccination in other countries [2,9-11]. Although information about TTS was disseminated by the U.S. media [30], differences in reporting rates could be associated with relative under-reporting or under-ascertainment in the United States, true differences in TTS risk after Ad26.COV2.S vs. ChAdOx1 nCoV-19 vaccination, differences in the populations vaccinated, or other factors.

A recent report estimated that the background incidence of cerebral venous sinus thrombosis with thrombocytopenia during the pre-COVID-19 era (2018-2019) [Amanda Payne, personal communication] was 0.16-0.22 cases per 100,000 persons per year. Based on these data, the expected 2-week incidence per million persons is 0.0062, an estimate similar to the reporting rate for TTS following mRNA COVID-19 vaccination (0.0057) found here. The small number of TTS cases reported following mRNA COVID-19 vaccination were among patients with different demographic characteristics (older, non-white males) and medical histories (e.g., presence of cardiovascular risk factors) than most cases reported following Ad26.COV2.S vaccination. These findings suggest that the TTS cases following mRNA COVID-19 vaccination represent a background rate of autoimmune HIT or TTS associated with a different risk factor than cases associated with Ad6.COV2.S vaccination.

Based on positive results from functional HIT platelet assays among early case reports, the pathophysiology of TTS following ChAdOx1 nCoV-19 vaccine receipt is believed to be similar to autoimmune HIT in which anti-PF4 antibodies activate platelets and cause thrombosis [1-3]. The binding site for anti-PF4 antibodies in patients with TTS reported after ChAdOx1 nCoV-19 vaccination is identical to the heparin binding site for HIT patients [31]. However, as was described with the initial 12 cases of cerebral venous sinus thrombosis with thrombocytopenia following Ad26.COV2.S vaccination, most TTS cases reported following Ad26.COV2.S vaccination for whom a serotonin release assay (the most common functional HIT platelet assay performed) was performed had negative results. Other investigators have described assay modifications needed for the serotonin release assay to identify patients with TTS following ChAdOx1 nCoV-19 vaccination [32] and the same modifications in testing procedure may be needed for TTS following Ad26.COV2.S vaccination.

Compared to the UK Expert Hematology Panel case definitions for “Definite VITT” or “Probable VITT” [17], the TTS case definition used here excludes cases without thrombocytopenia and can include cases with negative anti-PF4 antibody ELISA results, symptom onset less than 5 days after vaccination, and with normal D-dimer results. Our case definition appears to have greater sensitivity (Supplemental Results; *Comparison with UK Expert Hematology Panel case definition)* and may be more appropriate for a new condition in which the spectrum of disease is being actively investigated. However, consideration may be needed for inclusion of additional presentations such as positive anti-PF4 antibody ELISA lacking either thrombosis or thrombocytopenia or requiring cases following mRNA COVID-19 vaccination (or other non-adenoviral-vector COVID-19 vaccines in the future) to have positive anti-PF4 antibody ELISA results. Because evidence to date is insufficient to conclude that TTS is epidemiologically linked to receipt of mRNA COVID-19 vaccines, this requirement might limit false positives for the rare events that meet the TTS case definition following mRNA COVID-19 vaccination.

This report is subject to limitations. VAERS is a passive surveillance system, so the TTS reporting rates described likely underestimate true rates of occurrence. In addition, lack of anti-PF4 antibody ELISA testing could lead to both underreporting and underrepresentation of Tier 2 cases, compared to Tier 1 TTS cases. However, a strength of the current analysis is that because the COVID-19 immunization program was federally coordinated, all U.S. public health jurisdictions reported the number of COVID-19 vaccines that were administered, allowing a better estimate of reporting rates than usually possible when using VAERS data.

We estimate reporting rates of TTS following COVID-19 vaccination and describe the epidemiology of this emerging syndrome through a systematic surveillance effort. The epidemiology and hypothesized pathogenesis of TTS support a causal association with receipt of the Ad26.COV2.S COVID-19 vaccine. However, the much lower reported rates and difference in demographics for TTS cases following mRNA-based COVID-19 vaccines suggest that reports occurring after mRNA-based vaccine likely represents a background rate of cases of autoimmune HIT. Continued vigilance for TTS cases following Ad26.COV2.S vaccination is needed; the reporting rates and epidemiologic data about TTS following Ad26.COV2.S vaccination will be useful in formulating benefit-risk assessments for COVID-19 immunization programs in the United States and other countries [33-34].

## Supporting information

Supplemental Methods and Results

## Data Availability

Due to legal restrictions related to confidentiality, the original data are not available to the public.

## CDC disclaimer

The findings and conclusions in this report are those of the authors and do not necessarily represent the official position of the Centers for Disease Control and Prevention (CDC) or the Food and Drug Administration (FDA). Mention of a product or company name is for identification purposes only and does not constitute endorsement by the CDC and FDA.

## Acknowledgements

We would like to thank the following CDC staff who contributed to this manuscript without compensation aside from their salaries. For data collection: Suzanne Beavers, MD (CDC COVID-19 Response), Rebecca Byram, MS, MPH (CDC COVID-19 Response), Kathy Byrd, MD, MPH (CDC COVID-19 Response), Margaret Cortese, MD (CDC COVID-19 Response), Alice Guh, MD, MPH (CDC COVID-19 Response), Theresa A. Harrington, MD, MPH&TM (CDC COVID-19 Response), Amelia Jazwa, MSPH (CDC COVID-19 Response), Anamika Khatri-Dua, MD (CDC COVID-19 Response), Susan Lukacs, DO, MSPH (CDC COVID-19 Response), Michael M. McNeil, MD, MPH (CDC COVID-19 Response), Duong Nguyen, DO (CDC COVID-19 Response), Monica Parise, MD (CDC COVID-19 Response), Agam Rao (CDC COVID-19 Response), Allan Taylor, MD, MPH (CDC COVID-19 Response). For leadership and support: Denise Cardo, MD (CDC COVID-19 Response), Frank DeStefano (CDC COVID-19 Response). We would also like to thank the following individuals who contributed to this manuscript with funding support through the CISA Project. For programmatic support: Paula Campbell, MS, MPH (Vanderbilt University) and Braxton Hern, BS (Vanderbilt University). For vaccine safety expertise: Elizabeth Barnett, MD (Boston University), Anna Durbin, MD (Johns Hopkins University), Neal Halsey, MD (Johns Hopkins University), Nicola Klein, MD, PhD (Kaiser Permanente Northern California), Philip LaRussa M.D. (Columbia University), Stephen Pelton, MD (Boston University), Elizabeth Schlaudecker, MD, MPH (Cincinnati Children’s Hospital Medical Center), Michael Smith, MD, MSCE (Duke University), Mary Staat, MD, MPH (Cincinnati Children’s Hospital Medical Center), and Melissa Stockwell (Columbia University). We would also like to thank the clinical staff who have cared for these patients and who reported these events to VAERS.

