## Supplemental Methods and Results for "Case Series of Thrombosis with Thrombocytopenia Syndrome following COVID-19 vaccination—United States, December 2020–August 2021"

**VAERS review**

Potential reports of TTS were identified using combinations of searches, where each search was a group of selected PTs and/or text:

Search 1 searched for the words "thrombocytopenia" or "low platelets" in the text of the symptom or laboratory test fields, or for one or more of the following PTs: autoimmune heparin-induced thrombocytopenia, heparin-induced thrombocytopenia, immune thrombocytopenia, non-immune heparin associated thrombocytopenia, spontaneous heparin-induced thrombocytopenia syndrome, thrombocytopenia, thrombocytopenic purpura.

Search 2 searched for one or more of the following PTs: acute myocardial infarction, basal ganglia stroke, brain stem stroke, hemorrhagic stroke, hemorrhagic transformation stroke, ischemic stroke, lacunar stroke, myocardial infarction, perinatal stroke, silent myocardial infarction, spinal stroke, thrombotic stroke, vertebrobasilar stroke.

Search 3 searched for one or more of the following PTs: axillary vein thrombosis, deep vein thrombosis, pulmonary embolism.

Search 4 searched for one or more of the following PTs: aortic embolus, aortic thrombosis, aseptic cavernous sinus thrombosis, brachiocephalic vein thrombosis, brain stem embolism, brain stem thrombosis, carotid arterial embolus, carotid artery thrombosis, cavernous sinus thrombosis, cerebral artery thrombosis, cerebral venous sinus thrombosis, cerebral venous thrombosis, femoral artery embolism, hepatic artery embolism, hepatic artery thrombosis, hepatic vein embolism, hepatic vein thrombosis, iliac artery embolism, jugular vein embolism, jugular vein thrombosis, mesenteric artery embolism, mesenteric artery thrombosis, mesenteric vein thrombosis, obstetrical pulmonary embolism, portal vein embolism, portal vein thrombosis, portosplenomesenteric venous thrombosis, pulmonary artery thrombosis, pulmonary thrombosis, pulmonary venous thrombosis, renal artery thrombosis, renal embolism, renal vein embolism, renal vein thrombosis, splenic artery thrombosis, splenic embolism, splenic thrombosis, splenic vein thrombosis, spontaneous heparin-induced thrombocytopenia syndrome, subclavian artery embolism, subclavian vein thrombosis, superior sagittal sinus thrombosis, thrombosis mesenteric vessel, transverse sinus thrombosis, truncus celiacus thrombosis, vena cava embolism, vena cava thrombosis, visceral venous thrombosis. Search 5 searched for one or more of the following PTs: autoimmune heparin-induced thrombocytopenia, heparin-induced thrombocytopenia, heparin-induced thrombocytopenia test positive, idiopathic thrombocytopenic purpura, immune thrombocytopenia, immune thrombocytopenic purpura, non-immune heparin associated thrombocytopenia, platelet count decreased, severe fever with thrombocytopenia syndrome, spontaneous heparin-induced thrombocytopenia syndrome, thrombocytopenia, thrombocytopenic purpura, thrombotic thrombocytopenic purpura.

Search 6 searched for an extensive list of PTs describing a variety of conditions or procedures that could indicate thrombosis in a number of anatomic locations (available on request).

Potential reports of TTS were identified as reports containing PTs and/or text for both Search 1 and Search 2; both Search 1 and Search 3; both Search 1 and Search 4; or both Search 5 and Search 6. Reports identified by these preliminary searches were then reviewed by clinical abstractors. Reports with COVID-19 vaccine receipt date through August 31, 2021 were included in this review. Some cases were first identified through CDC's Clinical Immunization Safety Assessment (CISA) Project [1] when healthcare providers requested clinical consultation, and then subsequently reported to VAERS.

For reports that, upon initial review, appeared consistent with this case definition, an abstractor attempted to reach the best contact listed on the VAERS report form (typically the medical care provider for the reported patient), and medical records for the reported case were requested. Confirmation of case status was obtained through medical record review of the initial hospital course and/or discussions with treating clinicians. Clinical data about potential cases were reviewed with physician investigators from CDC's CISA Project [1], including specialists in infectious diseases, neurology, and hematology. If, after review, the clinical syndrome was not considered consistent with TTS, or if a more likely cause of the patient's signs and symptoms was identified, that report was removed from the case series. Specifically, we excluded cases with any of the following characteristics: active COVID-19 infection at the time of TTS diagnosis, pre-existing thrombocytopenia, TTS symptoms preceding COVID-19 vaccination, thrombocytopenia that was most likely artifactual, or thrombosis that was more likely due to other causes.

**Vaccination administration data**

Vaccine administration data were obtained for doses 1 and 2 of the Pfizer-BioNTech COVID-19 Vaccine and Moderna COVID-19 Vaccine, and dose 1 of Ad26.COV2.S from December 14-August 31, 2021 as reported to CDC from state health departments and other U.S. public health jurisdictions (e.g., U.S. territories) [2]. Dose administered data from Texas were available stratified by age group with one age group representing 25–39 year-olds; age group doses administered from Texas for age groups 25–29 and 30–39 years were estimated by applying to proportion of doses from other jurisdictions (i.e., other than Texas) among 25–39 year-olds that were among 25–29 year-olds and 30–39 year-olds to the Texas 25–39 year-old category. For other jurisdictions (i.e., other than Texas), doses with unknown sex or age (<1% of total) were excluded from calculation of reporting rates. For Texas, doses with unknown sex (0.5% of total) were excluded from calculation of reporting rates (no cases with unknown age were reported)

**Data analysis**

D-dimer values reported in D-dimer units were converted to fibrinogen equivalent units by multiplying by 2 [35]. Potential correlation between platelet nadir values and outcomes for TTS cases following Ad26.COV2.S was assessed through scatter plots; linear (Pearson) correlation between platelet nadir values and time from symptom onset to admission was also assessed. A public announcement in the United States of the existence of TTS cases following Ad26.COV2.S occurred on April 13, 2021 [7]; a pause in Ad26.COV2.S administration began and preliminary treatment guidance also was provided on the same date, including a recommendation to avoid use of heparin in patients with TTS. Treatment of patients admitted to hospital before the pause in use of Ad26.COV2.S was compared to treatment of patients admitted after the pause. [7]. Anti-PF4 antibody ELISA results and functional HIT platelet assay findings (e.g., serotonin release assay, latex immunoturbidimetric assay, P-selectin expression assay) were summarized [13].

**Supplemental results**

*Comparison with UK Expert Hematology Panel case definition*

Of the 50 TTS cases reported following Ad26.COV2.S, 29 (58%) were “Definite VITT,” 15 (30%) were “Probable VITT,” and 3 (6%) were “Possible VITT” using the UK Expert Hematology Panel case definition [3]. Three cases did not fall into any category using the UK case definition. These three all had cerebral venous sinus thrombosis, thrombocytopenia, but D-dimer <2.0 µg/mL FEU. One had a positive anti-PF4 antibody ELISA test, one did not have anti-PF4 antibody ELISA testing performed, and one had a negative anti-PF4 antibody ELISA test.

**Supplemental table 1:** Comparison of demographics, venous thromboembolism risk factors, and other selected characteristics for thrombosis with thrombocytopenia cases with cerebral venous sinus thrombosis (CVST) vs cases without CVST following receipt of Ad26.COV2.S (Janssen/Johnson & Johnson COVID-19 Vaccine)—United States, December 2020–August 2021

| | n (%) | | |
|---|---|---|---|
| **Characteristic** | **Total (N=50)** | **Cases with CVST (N=27)** | **Cases without CVST (N=23)** |
| Age in years, median (range) | 43.5 (18-70) | 44 (18-70) | 43 (24-65) |
| Female | 35 (70) | 22 (81) | 13 (57) |
| Race/ethnicity | | | |
| Hispanic | 3 (6) | 2 (7) | 1 (4) |
| White, non-Hispanic | 37 (74) | 19 (70) | 18 (78) |
| Black, non-Hispanic | 2 (4) | 1 (4) | 1 (4) |
| White, ethnicity not reported | 6 (12) | 5 (19) | 1 (4) |
| Unknown race and ethnicity | 2 (4) | 0 | 2 (9) |
| Venous thrombosis risk factors | | | |
| Obesity | 23 (46) | 14 (52) | 9 (39) |
| Hypertension | 14 (28) | 9 (33) | 5 (22) |
| Diabetes | 5 (10) | 3 (11) | 2 (9) |
| Systemic estrogen therapy* | 3 (6) | 2 (7) | 1 (4) |
| Other venous thrombosis risk factor† | 3 (6) | 1 (4) | 2 (9) |
| Pregnancy or postpartum (within 12 weeks) | 0 | 0 | 0 |
| Coagulation disorder | 0 | 0 | 0 |
| Time from vaccination to symptom onset in days, median (range) ‡ | 10 (0-18) | 8 (3-15) | 10 (0-18) |
| Time from symptom onset to hospital admission in days, median (range) ‡ | 5 (0-30) | 5 (0-30) | 5 (0-14) |
| Platelet nadir, median (range), cells/µL | 32,500 (5,000-127,000) | 33,000 (5,000-127,000) | 32,000 (6,000-112,000) |

Abbreviations: CVST, cerebral venous sinus thrombosis

* Systemic estrogen includes combined oral contraceptives (n=2) and an estradiol patch for hormone replacement (n=1)

† Other venous thrombosis risk factor: cirrhosis, malignancy, fertility treatment, venous catheter at thrombosis site (n=1 for each); one case had both malignancy and venous catheter at thrombosis site.

‡ Vaccine to symptom onset and symptom onset to admission: one case excluded because symptom onset date could not be determined

**Supplemental table 2:** Most common initial symptoms of patients with thrombosis with thrombocytopenia syndrome following receipt of Ad26.COV2.S (Janssen/Johnson & Johnson COVID-19 Vaccine)—United States, December 2020–August 2021

| **Symptom*** | **All patients, n (%) (N=50)** | **Patients with cerebral venous sinus thrombosis, n (%) (N=27)** | **Patients without cerebral venous sinus thrombosis, n (%) (N=23)** |
|---|---|---|---|
| Headache | 35 (70) | 24 (89) | 11 (48) |
| Nausea | 10 (20) | 7 (26) | 3 (13) |
| Fever | 9 (18) | 6 (22) | 3 (13) |
| Abdominal pain | 9 (18) | 5 (19) | 4 (17) |
| Chills | 9 (18) | 4 (15) | 5 (22) |
| Myalgias | 8 (16) | 4 (15) | 4 (17) |
| Fatigue, general malaise, or lethargy | 5 (10) | 2 (7) | 3 (13) |
| Vomiting | 6 (12) | 5 (19) | 1 (4) |
| Focal weakness | 6 (12) | 2 (7) | 43 (17) |
| Photophobia | 4 (8) | 3 (11) | 1 (4) |
| Bruising | 3 (6) | 1 (4) | 2 (9) |
| Cough† | 3 (6) | 2 (7) | 1 (4) |
| Dizziness or lightheadedness | 3 (6) | 1 (4) | 2 (9) |
| Extremity swelling | 3 (6) | 0 | 3 (13) |
| Extremity pain | 4 (8) | 0 | 4 (17) |
| Visual disturbance | 3 (6) | 3 (11) | 0 |
| Back pain | 3 (6) | 1 (4) | 2 (9) |
| Chest pain | 2 (4) | 2 (7) | 0 |
| Neck pain | 2 (4) | 2 (7) | 0 |
| Shortness of breath | 2 (4) | 1 (4) | 1 (4) |
| Facial droop | 2 (4) | 1 (4) | 1 (4) |

* Table includes initial symptoms that were present in at least 2 patients; symptoms present in only one patient included anorexia, diarrhea, difficulty thinking, digital pain, ear pain, joint pain, numbness, petechiae, rash, rectal bleeding, seizure, sinus pressure, toe pain

† Includes one patient with hemoptysis

**Supplemental figure 1:** Distribution of hospital length of stay (in days) for thrombosis with thrombocytopenia syndrome cases following receipt of Ad26.COV2.S (Janssen/Johnson & Johnson COVID-19 Vaccine)—United States, December 2020–August 2021, for patients discharged alive (N=44).

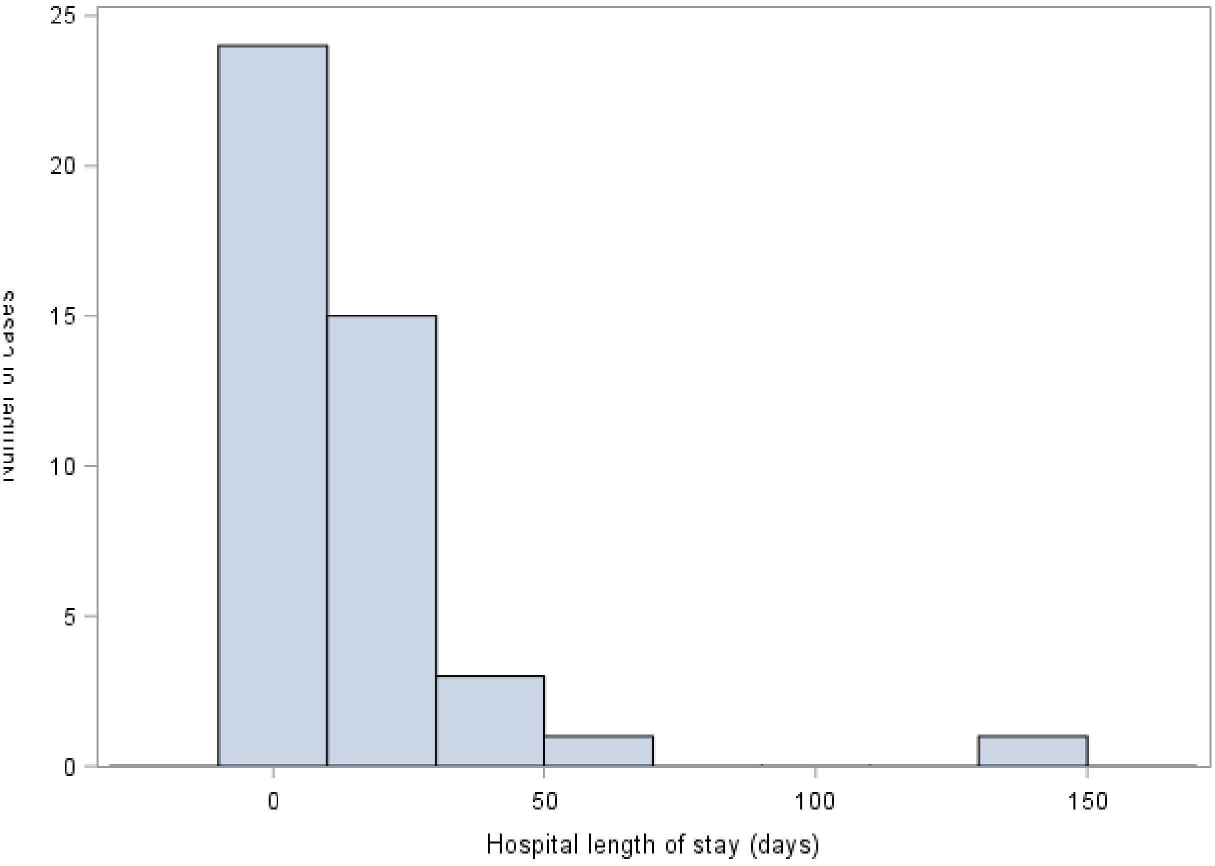

**Supplemental figure 2:** Distribution of days from vaccination to symptom onset for thrombosis with thrombocytopenia syndrome after receipt of Ad26.COV2.S (Janssen/Johnson & Johnson COVID-19 Vaccine)—United States, December 2020–August 2021 (N=49). Excludes one patient whose symptom onset date could not be determined.

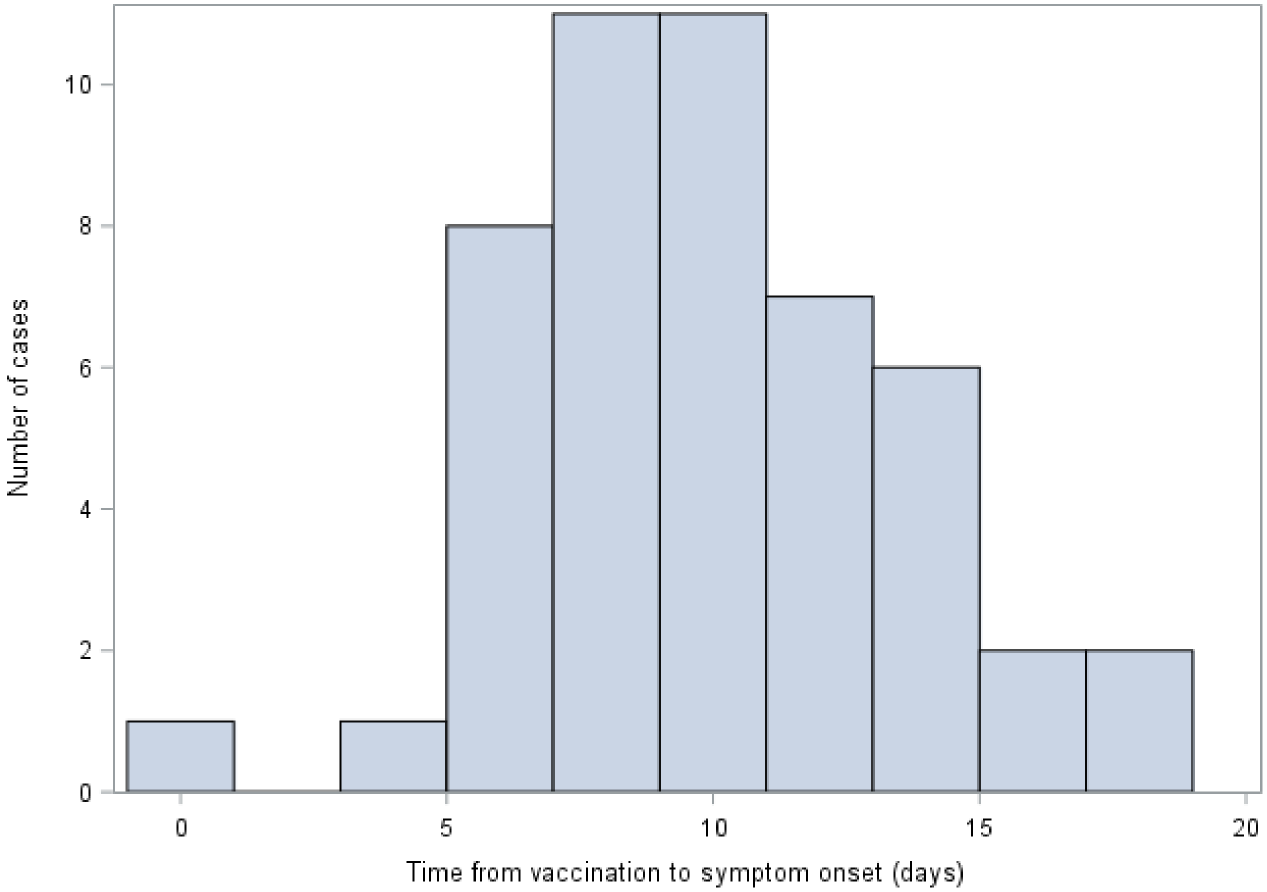

**Supplemental figure 3:** Distribution of platelet nadir counts for thrombosis with thrombocytopenia syndrome cases following receipt of Ad26.COV2.S (Janssen/Johnson & Johnson COVID-19 Vaccine)—United States, December 2020–August 2021

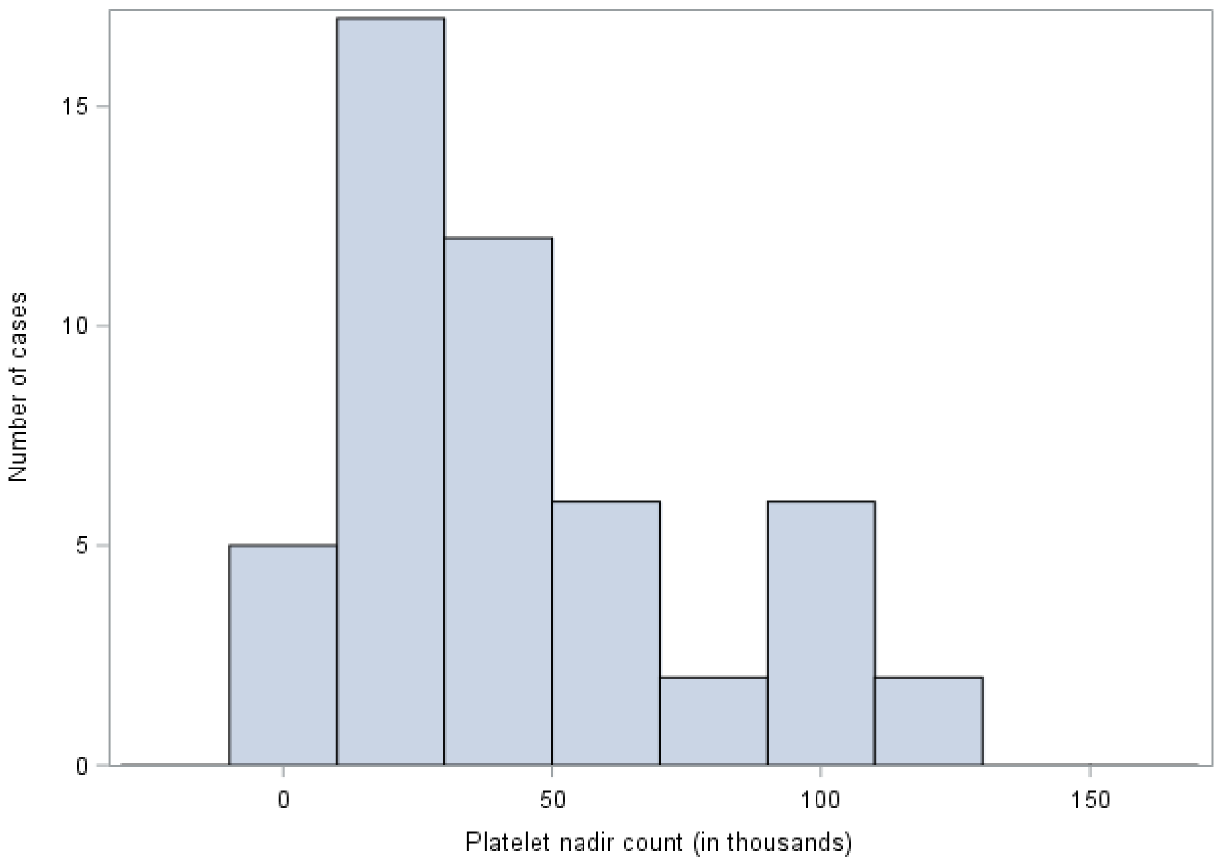

**Supplemental figure 4:** Scatter plot of platelet nadir count versus hospital length of stay (in days) for thrombosis with thrombocytopenia syndrome cases following receipt of Ad26.COV2.S (Janssen/Johnson & Johnson COVID-19 Vaccine)—United States, December 2020–August 2021 (N=44). Limited to patients who were discharged from the hospital alive.

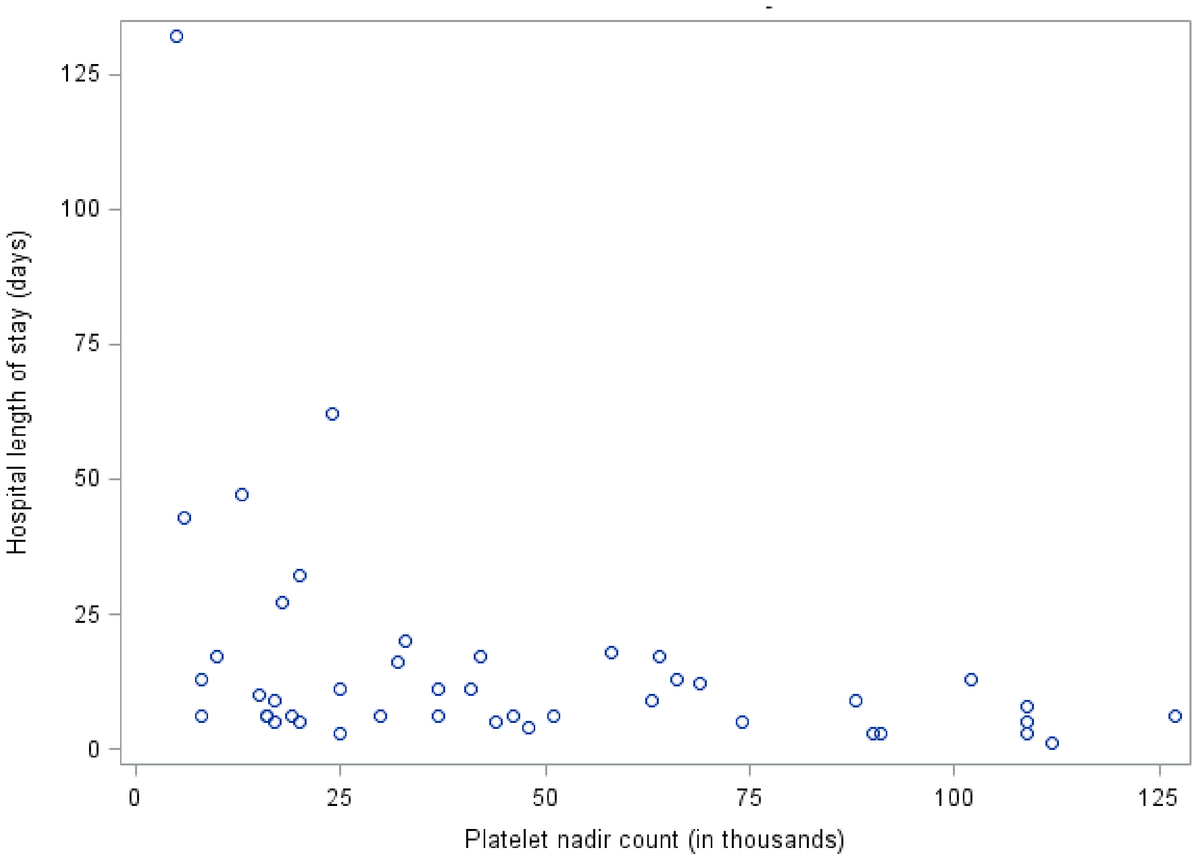

**Supplemental figure 5:** Scatter plot of platelet nadir value vs time (in days) from symptom onset to hospital admission for thrombosis with thrombocytopenia syndrome cases following receipt of Ad26.COV2.S (Janssen/Johnson & Johnson COVID-19 Vaccine)—United States, December 2020–August 2021 (N=49). Excludes one patient whose symptom onset date could not be determined. $R^2$ value = 0.25 (P=0.003).

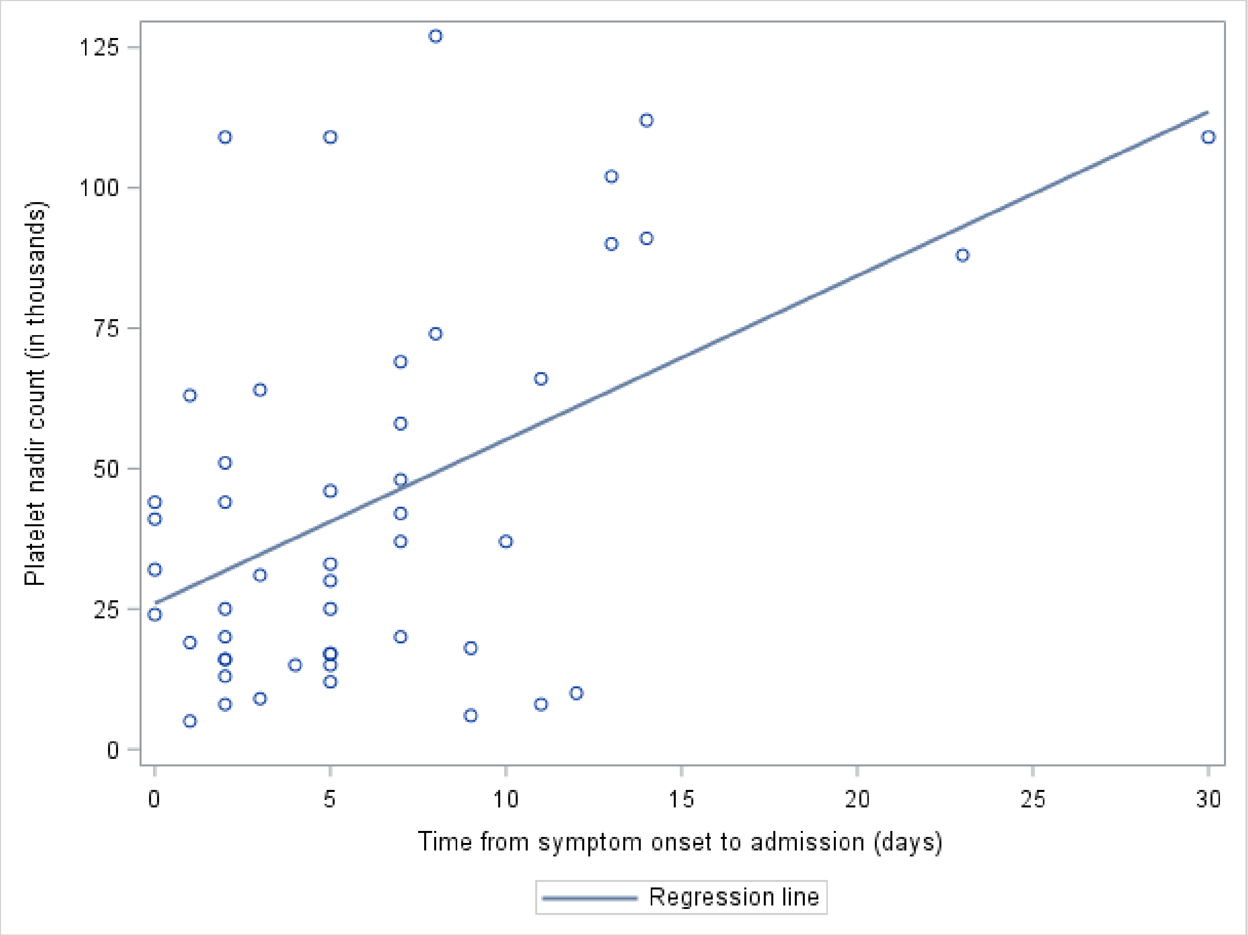
